# Understanding Clinician Edits to Ambient AI Draft Notes: A Feasibility Analysis Using Large Language Models

**DOI:** 10.64898/2026.02.27.26347290

**Authors:** Yawen Guo, Yiliang Zhou, Di Hu, Sairam Sutari, Emilie Chow, Steven Tam, Danielle Perret, Deepti Pandita, Kai Zheng

## Abstract

Ambient AI documentation tools generate draft notes that clinicians can review and edit before signing off in electronic health records. Scalable computational approaches to characterize how clinicians modify drafts remain limited, yet are essential for evaluating and improving AI effectiveness. We examined the feasibility of a few-shot prompted large language model (LLM) for categorizing sentence-level edits between AI drafts and final documentation. We developed five label-specific binary models targeting medication, symptom, diagnosis, orders/tests/procedures, and social history edits, and refined prompts using adversarial negatives and verification gates. Evaluation was performed against a human-annotated corpus. Medication and symptom models achieved promising performance (F1=0.787 and 0.780), whereas remaining models were precision-limited. Errors clustered in long, complex edits and category-boundary ambiguity. Therefore, prompt engineering is reliable for categorizing edits with explicit clues, while for complex context-dependent categories they are better suited for triage by labeling edits for human review.

## Introduction

Ambient artificial intelligence (AI) documentation systems can draft clinical notes by transcribing and summarizing clinical visit audio, shifting documentation into a workflow in which clinicians review and revise AI-generated content before committing into electronic health record (EHR) systems ^1–4^. Prior studies have emphasized benefits such as reduced documentation burden, improved efficiency, and favorable user experience ^5–8^. However, these evaluation metrics offer limited visibility into where AI drafts fall short. The edits clinicians make to AI drafts can provide a direct lens on system reliability and usefulness by revealing what the draft omitted, or framed in ways that clinicians judged insufficient for clinical communication and note quality. In our prior content-analysis study, we compared ambient AI-drafted notes with clinician-finalized versions, derived sentence-level edit units that retain local context to interpret each modification, and manually annotated each sentence unit of analysis with an editing category taxonomy ^9,10^. This framework supports fine-grained characterization of what clinicians change, but extending it to routine deployment remains challenging.

Manual annotation is resource-intensive, making it difficult to quantify how edit patterns vary across specialties and note sections ^11^. Traditional natural language processing (NLP) classifiers also face practical and methodological barriers in this setting since edited content is often heterogeneous, with multiple changes spanning different clinical concepts. For example, diagnosis-related edits may hinge on subtle framing or implied reasoning that is difficult to capture with keyword-based signals. In addition, supervised classifiers typically require substantial task-specific training data and ongoing maintenance as documentation templates, clinical language, and ambient AI outputs evolve, limiting their usefulness for rapid, iterative measurement in real-world deployments ^12^. Large language models (LLMs) prompting offer a pragmatic path toward scaling edit categorization because these labels depend on clinical meaning rather than surface similarity alone (e.g., lexical overlap or edit distance) ^13–15^. In practice, feasibility of prompt engineering depends on how explicitly the target information is expressed in the local text unit and the amount of clinical context required to interpret it. For instance, medication edits often contain explicit cues such as drug names and regimen attributes, whereas diagnosis-related edits may be subtle, context-dependent, and difficult to separate from symptom descriptions or narrative reframing ^16^.

This paper evaluates the feasibility of using few-shot prompting LLMs to categorize clinicians’ edits to AI drafts using sentence-level edit units derived from paired draft–final notes. Building on our prior edit unit cohort and label reference annotations, we reframe the task as five label-specific binary models (medications, symptoms, diagnoses, orders/procedures, and social context), each required to output a structured decision with evidence content from the *before* and *after* note text ^9^. We designed a prompt refinement strategy for performance control and auditability, including adversarial negatives and an evidence verification gate. We operationalize feasibility in terms of (i) categorization performance (precision, recall, and F1) and (ii) operational output completion rate under runtime constraints. Importantly, we evaluate feasibility under realistic deployment constraints for protected health information (PHI) including limited GPU resources under HIPAA-compliant environments, and restricted use of external APIs or additional training levers. Our goal is to identify which edit categories support high-precision automated monitoring versus those better suited for high-recall candidate retrieval with targeted human review.

## Methodology and Materials

### Data and Task Formulation

We used data from University of California Health, where two commercial ambient AI systems were piloted in outpatient settings from late 2023 through mid 2025. The source data include ambient AI generated draft note sections and the corresponding clinician finalized note sections. Clinicians can selectively choose among four note sections to insert into the notes and review, including History of Present Illness, Physical Exam, Assessment & Plan, and Results. This feasibility study built on the paired draft–final section dataset and sentence-level edit units developed in our prior content-analysis study ^9^. In that work, we sampled notes to represent various specialty and note sections, and aligned AI-drafted and clinician-finalized note sections to derive sentence-level edit units that retain sufficient surrounding context to interpret each change. The sentence-level edit units served as the unit of analysis for this study and were constructed to be non-overlapping within a section by merging adjacent/overlapping text spans into a single contiguous unit. Each unit of analysis consists of a *Before* span from the AI draft and an *After* span from the clinician-final text. The resulting gold-standard corpus comprised 313 draft–final note section pairs from 200 clinical encounters, and 713 sentence-level edit units. Full details of cohort construction, edit-unit derivation, and manual annotation procedures are reported in the prior manuscript ^9^.

In this feasibility study, we operationalized edit classification as a set of label-specific binary categorization tasks at the edit-unit level. Each model evaluated whether a target edit category was present in the *Before* and *After* text for a given edit unit. We focused on five clinically meaningful categories identified that are routinely documented across outpatient encounters and are common targets of clinician revision: medication-related edits (E-Med), symptom-related edits (E-Sym), diagnosis-related edits (E-Dx), test/order/procedure-related edits (E-Test), and social-context edits (E-Soc). We separated the annotated edit units into three non-overlapping subsets: (1) a training set (n=313) used only to select few-shot examples and draft initial prompts, (2) a development set (n = 200) used for iterative prompt refinement and error analysis, and (3) a held-out test set (n = 200) that was frozen and used only for final evaluation and reporting performance. We performed this random split once on the full annotated edit-unit dataset and reused the same partitions for all five models. Edit category definitions and illustrative de-identified examples are summarized in Table 1. This study is approved by University of California, Irvine Institutional Review Board (UCI IRB #7123).

**Table 1.**
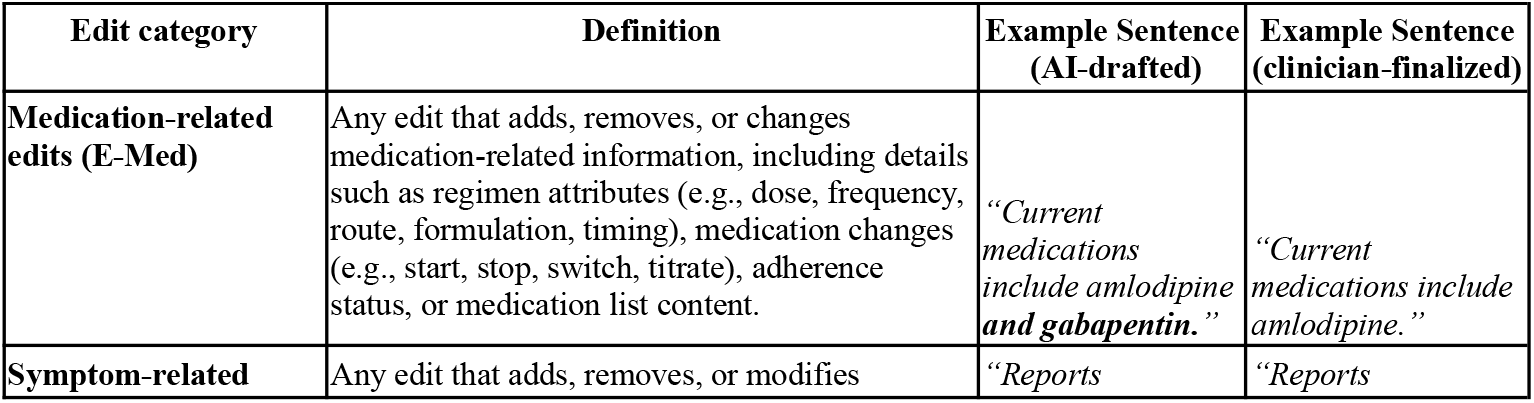
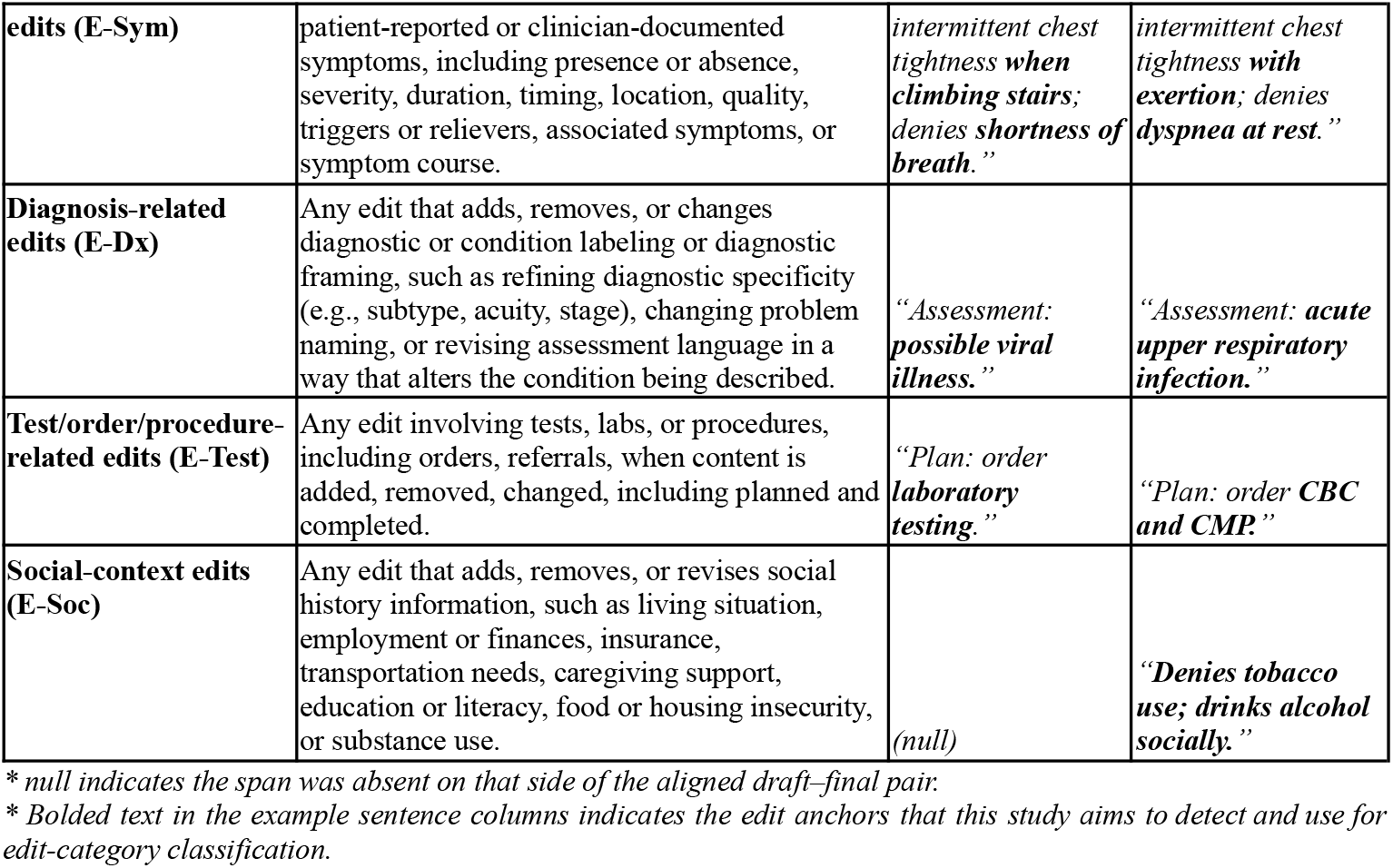
Edit category definitions and de-identified *Before/After* examples for binary classification.

### LLM inference and prompt design

We conducted a few-shot, prompt-based inference using the open-weight instruction-tuned model meta-llama/Llama-3.2-3B-Instruct implemented with Hugging Face Transformers ^17,18^. All experiments were run on a single NVIDIA T4G GPU (16 GB VRAM) with NVIDIA driver version 580.95.05 and CUDA 13.0. Inference was performed with PyTorch 2.9.1 and Hugging Face Transformers 4.57.6 (Accelerate 1.12.0) using the standard *model*.*generate()* API. We used deterministic decoding (do_sample=False) with max_new_tokens=100 and a per-request time limit of 30 seconds. Prompts were processed one edit unit per request (batch size = 1). All data processing and model inference were conducted within a HIPAA-compliant, institution-approved AWS environment with appropriate access controls. A system message specified the target edit category and required a structured JSON output, and a user message provided metadata (note ID, section name, edit-unit order) along with the edit-unit text ^19^. Edit-unit order is a within-section sequence number used to uniquely index multiple edit units within the same note section.

Prompts were refined iteratively on the development set through error analysis, including adding targeted adversarial negative examples reflecting common confusions for each category and introducing a brief structured self-check that required the model to apply inclusion and exclusion criteria before deciding ^20,21^. In addition, we implemented a prompt-level verification gate by requiring that positive predictions be supported by qualifying, verbatim evidence excerpts from the edit unit. Specifically, the prompt instructed the model to output *present=true* only when it could quote category-appropriate “anchor” evidence (e.g., medication anchors for medication-related edits, diagnostic labels for diagnosis-related edits) and to output *present=false* with evidence marked as N/A otherwise. During the development stage, we also experimented with prompts that requested stepwise rationale; the final prompts retained only the structured self-check and the evidence-based output constraints, without collecting reasoning details ^22^. For each model, we used a set of few-shot examples consisting of approximately 4–6 positives and 18–20 targeted negatives; the composition of examples was adjusted during development to address observed error patterns, and then frozen for the held-out test evaluation. All models shared the same prompt template and JSON output schema, with category-specific components limited to the operational definition, boundary/anchor rules, and examples. We summarize prompt structure in Figure 1. Full prompts are available from the first author upon reasonable request.

**Figure 1.**
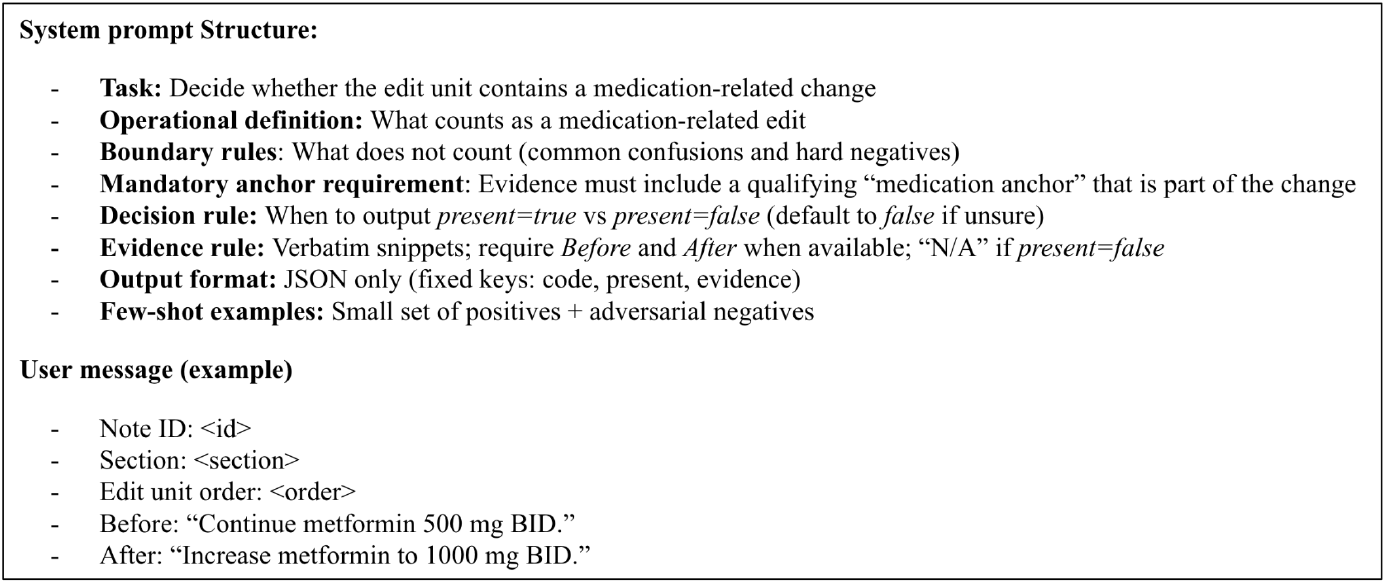
Prompt structure for edit classification (example: medication-related edits).

### Structured outputs, post-processing, and evaluation

Models were instructed to return a single JSON object containing a binary decision and a short evidence field with verbatim excerpts from the edit unit when *present=true*. Because parameter-efficient model generations might produce malformed JSON, we implemented a tolerant parser that extracts the first JSON-like object from the response and supports lightweight normalization (e.g., trimming non-JSON prefixes/suffixes and correcting minor quoting/punctuation issues) before parsing. In the final held-out evaluation, all outputs were valid JSON and no further edits were required. Instances that produced no output due to runtime limits or other execution errors were logged and excluded from metric computation. Prompt refinement was completed before running the held-out test set. Final performance was assessed on the held-out set of 200 edit units. We report label-specific precision, recall, and F1 on instances with successfully parsed outputs. We manually reviewed false positives (FPs) and false negatives (FNs) for each best-performing model and summarized recurring failure modes. Figure 2 summarizes the end-to-end workflow.

**Figure 2.**
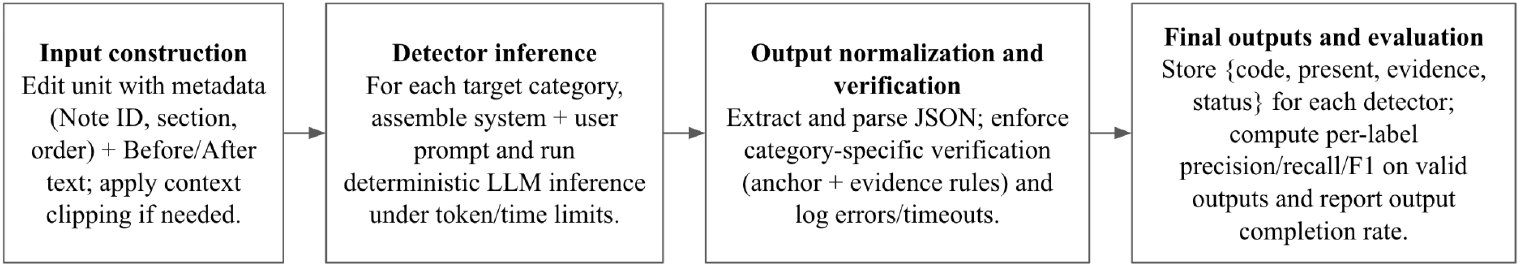
End-to-end workflow for LLM-based edit-category classification and verification.

## Results

We evaluated few-shot prompting for five edit types using an iterative development protocol on a development set, followed by a locked evaluation on a held-out test set. We present results in three parts. First, we use E-Med as an illustrative case study to show how targeted prompt refinements improved precision and recall over successive iterations. Second, we report held-out test performance for the best-performing model for each edit type. Third, we characterize dominant failure modes through qualitative error analysis, highlighting where the prompt-only model falls short.

### E-Med prompt refinement case study

We curated few-shot prompt examples from the training set and iteratively evaluated prompt variants on a held-out development set (n=200) that was not used to construct the examples. After each run, we revised the prompt to address the dominant error pattern observed in the prior iteration. Starting from a definition-only zero-shot baseline, the model showed recall that was markedly low. Adding a small set of labeled examples shifted the model toward the intended decision boundary, substantially improving recall. However, expanding example coverage further increased sensitivity at the expense of over-triggering, causing a precision drop driven by medication-related language that did not meet the definition. To restore specificity, we introduced an explicit “exclusion” section plus adversarial negatives targeting the most frequent false-positive patterns. Finally, we added a mandatory pre-output verification gate requiring (i) an explicit medication-related anchor and (ii) evidence that is consistent with the edited span before allowing *present=true*. As summarized in Table 2, these successive refinements improved overall development-set performance, increasing F1 from 0.400 in the zero-shot baseline to 0.800 in the final prompting.

**Table 2.** E-Med prompt refinement progression (detailed key changes)

| Prompt version | Key changes to E-Med prompt | Precision | Recall | F1 |
| --- | --- | --- | --- | --- |
| v0 (zero-shot) | Definition-only baseline prompt:<br>Task framing and E-Med definition.<br>Inclusion/exclusion guidance without any labeled examples; no explicit checklist. | 0.667 | 0.286 | 0.4 |
| v1 | Introduce labeled examples to calibrate decision boundaries:<br>Added 1 positive and 4 negative examples from the training pool to reduce under-detection by grounding the abstract definition in concrete patterns. | 0.5 | 0.643 | 0.563 |
| v2 | Increase coverage of medication-change patterns:<br>Expanded to 4 positive and 8 negative examples to cover more surface forms (e.g., start/stop, dose change, frequency change, adherence/titration language, med list edits). | 0.519 | 1 | 0.683 |
| v3 | Target dominant false positives with explicit “what does NOT count.”:<br>Added a structured exclusion section derived from observed FPs (e.g., allergy-only mentions, problem/diagnosis changes without med action, labs/vitals without med change, generic “meds reviewed” without specific change).<br>Added 10 more adversarial negative examples that look medication-adjacent but should be *present=false*. | 0.632 | 0.857 | 0.727 |
| v4 | Add mandatory pre-output verification gate (rule-based checklist). *Before* allowing *present=true*, the model must confirm:<br>Explicit medication anchor exists (e.g., medication name, dose, frequency, route, formulation, titration, or adherence).<br>The anchor is part of the edit change (added/removed/modified between “*Before*” and “*After*”).<br>Evidence snippets contain only medication-related text. | 0.769 | 0.833 | 0.8 |
| **Final (locked test)** | Frozen v4 prompt ran on the held-out set. | 0.774 | 0.8 | 0.787 |

### Best-performing prompt-based model performance on the held-out test set

Table 3 summarizes performance for the best-performing model for each edit type on the held-out test set (n=200). Overall, the strongest performance was observed for E-Med (Precision=0.774, Recall=0.800, F1=0.787) and E-Sym (Precision=0.657, Recall=0.959, F1=0.780). E-Dx achieved moderate performance (Precision=0.560, Recall=0.836, F1=0.671). In contrast, E-Test (Precision=0.523, Recall=0.831, F1=0.642) and E-Soc (Precision=0.483, Recall=0.933, F1=0.636) showed lower precision despite high recall, indicating that these categories remain more challenging for prompt-only classification and are more prone to FPs in fully automated use.

**Table 3.** Best-performing model performance by edit type on the held-out test set.

| Edit Category | N_TP | N_FP | N_FN | N_TN | Prevalence (gold positives) | N failed (out-of-memory) | Precision | Recall | F1 |
| --- | --- | --- | --- | --- | --- | --- | --- | --- | --- |
| E-Med | 72 | 21 | 18 | 89 | 45.00% | 0 | 0.774 | 0.8 | 0.787 |
| E-Sym | 71 | 37 | 3 | 89 | 37.00% | 0 | 0.657 | 0.959 | 0.780 |
| E-Dx | 56 | 44 | 11 | 88 | 33.50% | 1 | 0.560 | 0.836 | 0.671 |
| E-Test | 69 | 63 | 14 | 52 | 41.50% | 2 | 0.523 | 0.831 | 0.642 |
| E-Soc | 56 | 60 | 4 | 80 | 30.00% | 0 | 0.483 | 0.933 | 0.636 |

\* TP = true positives; FP = false positives; TN = true negatives; FN = false negatives. Each edit type was evaluated on up to 200 test edit units. TP+FP+TN+FN may be <200 when some units encountered run-time errors and were excluded from scoring.
\* Prevalence is reported relative to the full test set. Other metrics are computed over the successfully processed units.

### Error patterns and boundary failures in prompt-based edit classification

To investigate where prompt-based models succeed and fail, we reviewed FPs and FNs for the best-performing model in each edit category (Table 4). Across models, the dominant FP pattern reflects boundary ambiguity in clinical language. This was most prominent among categories including symptoms, diagnoses, and orders/tests/treatment plans. Edits in assessment and problem-list text or care-plan statements were frequently “pulled” into the wrong category when explicit anchors were weak. For example, the symptom classification model tended to over-trigger on problem-list or plan phrasing that implies a symptom context but does not directly state a symptom change, while diagnosis and test models were prone to over-interpreting management-oriented wording (medications, procedures, referrals) as diagnostic or test changes. E-Soc model results showed a similar pattern when general clinical narrative or planning language was interpreted as social determinants of health (SDOH) in the absence of a clear social history anchor.

**Table 4.** Summary of model performance and dominant error modes by edit category.

| Edit Category | Dominant error direction | Top FP modes (n)* | Top FN modes (n)* | Key drivers |
| --- | --- | --- | --- | --- |
| E-Med | Balanced | Procedure/injection or follow-up language mis-triggering (5); diagnosis/differential wording mis-triggering (4); in long mixed edit units, evidence pulls non-target content (4). | Delete-only medication statements missed (8); medication embedded in long mixed content missed(6); implicit/uncertain medication decision language missed (e.g., “agents discussed...may reconsider”) (4). | FNs occur in delete-only or long mixed procedural blocks, and with implicit/generic medication decisions. Residual FPs reflect boundary bleed into procedure/diagnosis wording and occasional evidence-span drift. |
| E-Sym | Precision-limited | Diagnosis/assessment/problem-list edits mis-triggering (10); treatment/medication/procedure/care-plan language mis-triggering (10); non-symptom clinical history (e.g., “utilizing an assistive device...”) mis-triggering (6). | Symptom content embedded in surgical/history narrative missed (2). | Boundary overlap (symptoms vs diagnoses/treatments) drives mislabeling when the edited text does not explicitly mention a patient symptom or symptom denial, especially in template-like lists or delete-only units. |
| E-Dx | Precision-limited | Treatment/medication/procedure/surgery language mis-triggering (12); symptom-only or laterality/narrative updates mis-triggering (8); test/imaging/workup statements mis-triggering (6). | Temporality updates to prior diagnoses/illness history missed (4); delete-only diagnosis statements removed missed (4); diagnosis cues embedded in long exam/history expansions missed (2). | Diagnosis is tightly coupled with symptoms, tests, and management; single edit units often lack context to separate “diagnostic framing” from workup/plan language. Performance degrades on delete-only and long mixed-content edits. |
| E-Test | Precision-limited | Medication edits mis-triggering (15); Dx/assessment rewording mis-triggering (9); non-order narrative edits (10). | Referrals/therapy/follow-up actions missed (6); procedure/surgery/pre-op items missed (3); surveillance/awaiting results missed (2). | Boundary overlap (tests vs medication/diagnosis) and limited temporality cues (planned vs performed vs results). Delete-only edits reduce available evidence, amplifying misses and spurious triggers. |
| E-Soc | Precision-limited | Clinical diagnosis/assessment/problem-list edits mis-triggering (e.g., “Her anemia has worsened slightly”; “Venous insufficiency, possible calciphylaxis”) (16); care-plan / workup language (referral/PT/surgery planning/tests) mis-triggering (8); minor clinical narrative/date/laterality edits mis-triggering (4). | Obvious “social history” additions missed in long *before* and *after* context. (3). | Over-broad trigger space and weak SDOH anchor enforcement. Occasional evidence-text misalignment (pipeline/merge) can further inflate false positives. |

\* Top error modes shown; remaining errors were heterogeneous.

Long, complex edit units and template-like multi-item lists within a unit are associated with a consistent failure mode across categories: true anchors were present but buried among other problems, plans, or list items, and the model either missed them (FN) or extracted an overly broad evidence span that included nearby text that was not part of the target change (FP). This pattern was most visible in care-plan style units that bundle multiple actions, counseling statements, and follow-up instructions. In these blocks, the model may detect that a label-specific anchor is *present*, but it often cannot localize the specific edited subspan, leading to overly broad or misaligned evidence. Across diagnosis- and test-related edits, errors clustered when temporality or status determined the label. In many cases, the edit unit did not explicitly indicate whether something was planned, performed, resulted, or part of prior history. When temporality cues were implicit, models tended to either miss the change (FN) or misattribute it to related category (FP), especially when the text is written in a compressed “plan” style that blends diagnostic framing, workup, and management. Another cross-category error pattern is when the edit is delete-only or otherwise short and fractured. For example, in delete-only units where the *After* text is empty, the model may be left with only a single short line (e.g., “Continue vitamin D supplementation.”). In these cases, the model may infer the label from a minimal span without surrounding narrative, making it harder to distinguish content type.

## Discussion

Scaling edit classification between ambient AI drafts and clinician-final notes is essential for understanding how clinicians refine AI-generated documentation and where these AI systems fall short ^23^. It can support continuous monitoring, guide targeted model improvements, and ultimately inform safer and more efficient workflows. In this feasibility study, we evaluated few-shot prompting models using Llama-3.2-3B-Instruct, a 3B-parameter instruction-tuned large language model, deployed in a HIPAA-compliant environment with limited computing power and without additional labeled training data or EHR context ^24^. We found that prompt design choices, especially iteratively adding adversarial negative examples and applying an explicit verification gate before output, helped manage precision–recall tradeoffs and improved performance. The models performed best for edit types with clear, text-anchored signals, especially medication- and symptom-related edits. Performance was substantially weaker for more context-dependent categories such as social history, diagnosis, and test/order/procedure-related edits, indicating that prompt-only methods are not equally feasible across edit types and that some categories will require richer context or human review to support reliable downstream use.

These differences have direct implications for what can be used safely in practice. For categories with stronger performance, the most defensible near-term use is high-precision labeling to support scalable measurement and targeted quality monitoring ^25^. To avoid over-reliance on noisy outputs in future deployments, such use should be paired with safeguards such as confidence-thresholded acceptance, explicit reporting of coverage, and abstention for ambiguous edit units such as delete-only changes or long mixed-content spans ^26^. Feasibility should therefore be judged by whether the model can achieve sufficiently high precision at the coverage needed for the intended workflow, not by overall F1 alone. In contrast, the context-dependent categories that remained precision-limited in our evaluation should not be used for fully automated evaluation because false positives can systematically distort downstream estimates ^27^. Despite the labor required, manual review remains the most reliable way to characterize how ambient AI drafts perform and how clinicians revise them, and it is essential for establishing a trustworthy reference for evaluation. Until additional controls are introduced, these models are better positioned for triage workflows that surface candidate edits for targeted human review ^28^. From an operational perspective, the value of these labels also depends on the balance between time saved and time spent verifying outputs; triage deployments should therefore report or plan for the associated workload ^29^.

Failure cases in this study are concentrated in edit units where the available evidence is inherently weak, especially delete-only changes where the final text is missing and long mixed-content spans where multiple clinical concepts are interrelated. These boundary conditions suggest clear design implications for prompt-only classification at scale. One path is to develop prompts for known difficult structures, such as list-like templates, and for ambiguous contents of the note, such as distinguishing planned tests from performed procedures and requiring explicit anchor spans before accepting a positive prediction ^30^. A complementary path is to move beyond prompt-only methods via parameter-efficient fine-tuning on labeled edit units, particularly for cases that rely on implicit clinical reasoning ^31^. These adjustments can target the specific situations where edit unit context is least informative and where precision failures are most costly for downstream use ^27^.

We used a fixed prompt design to reflect realistic governance and deployment constraints; however, retrieval-based prompting may improve borderline, context-sensitive categories with high anchor variability, without requiring additional labeled training data ^32^. This direction is most relevant for complex edits, where dynamically retrieving label-specific examples could provide more appropriate reference cases than a fixed example set. At the same time, we used a 3B-parameter instruction-tuned large language model in a HIPAA-compliant environment with limited GPU memory, which contributed to out-of-memory failures and constrained prompt length and runtime. In the final prompt iterations, inference approached roughly 30 seconds per edit unit as prompts grew longer. This underscores a practical tradeoff in prompt engineering where adding detailed instructions, verification gates, and an expanded list of examples can improve performance but reduces throughput and scalability. Future work should therefore evaluate larger LLMs with stronger reasoning capability, redesign the task with hierarchical labels for vague boundaries such as separating test orders from procedures, and consider domain knowledge adaptation such as SDOH ontology where richer context may be necessary ^33,34^. In addition, lightweight rule-based components such as dictionary matching may further improve robustness when edit units mix multiple clinical concepts ^35^. Finally, scaling to tens of thousands of note sections will likely require more efficient inference strategies such as batching and optimized serving. Overall, our findings suggest that prompt-only classification is most feasible for edit types with explicit textual anchors, while context-dependent categories require further controls or added context before they can support reliable automated measurement.

## Conclusion

This feasibility study shows that few-shot, prompt-based LLM classifiers have the potential to scale edit-type categorization of clinician modifications to ambient AI draft notes, but performance varies substantially by content category. Under privacy and deployment constraints, models on medication- and symptom-related edits achieved strong held-out performance after iterative prompt refinement with hard negatives and an evidence-based anchor verification gate. In contrast, diagnosis, order/test/procedure, and social history classifiers remained precision-limited because sentence-level edits often blend clinically related content and key cues, and rationale are frequently implicit. These findings support a practical workflow: edits with explicit textual cues can be monitored automatically, whereas categories that typically require broader clinical context are better suited for labeling likely edit types for targeted human review. This work demonstrates a deployable pathway for scalable, auditable edit detection and classification that can help health systems and ambient AI vendors monitor common revision patterns in AI drafts, prioritize quality improvement targets, and evaluate changes over time. Future work should test larger LLMs with parameter-efficient fine-tuning, add lightweight rule-based signals, and redesign labels to support hierarchical drill down categorization for mixed content edits.

## Data Availability

The data contains protected health information (PHI) thus will not be available for share.

## Notes

### Competing Interest Statement

The authors have declared no competing interest.

### Funding Statement

This study did not receive any funding.

### Author Declarations

University of California, Irvine Institutional Review Board IRB #7123

